# Risk Factors for Hypertension in Indonesian Hajj Pilgrims: A Systematic Review and Meta-Analysis

**DOI:** 10.1101/2025.07.01.25330661

**Authors:** Muhammad Abdillah Roikhan, Faza Saiqur Rahmah

## Abstract

**Background:** Hypertension is a leading cause of morbidity and mortality among Indonesian Hajj pilgrims, a large and high-risk population with many elderly individuals. This study aims to quantitatively synthesize evidence on risk factors for hypertension in this specific population.

**Methods:** Following a pre-registered protocol (PROSPERO: CRD420251084054), a systematic literature search was conducted in major international and national databases up to June 2025. This review included analytical observational studies. Two reviewers independently performed study selection, data extraction, and risk of bias assessment (JBI checklist). A random-effects meta-analysis was performed, and the certainty of evidence was assessed using the GRADE framework.

**Results:** Five cross-sectional studies involving 286,987 pilgrims were included. The meta-analysis identified several significant risk factors for hypertension: advanced age, a history of Diabetes Mellitus (pooled OR: 1.86; 95% CI [1.72, 2.01]; I^2^=0%), obesity/excess BMI (pooled OR: 1.39; 95% CI [1.26, 1.53]; I^2^=34%), and a family history of hypertension (pooled OR: 1.70; 95% CI [1.29, 2.23]; I^2^=92%). Dyslipidemia was not found to be a statistically significant risk factor (pooled OR: 1.18; 95% CI [0.99, 1.40]; I^2^=94%). The certainty of evidence was rated low to moderate.

**Conclusion:** Advanced age, Diabetes Mellitus, obesity, and family history are major risk factors for hypertension in Indonesian Hajj pilgrims. These findings support a policy shift towards long-term health coaching and proactive screening during the extensive Hajj waiting period to improve pilgrim health outcomes.

**STRENGTHS AND LIMITATIONS OF THIS STUDY:**

- This is the first systematic review and meta-analysis to quantitatively synthesize evidence on hypertension risk factors specifically for the large and unique population of Indonesian Hajj pilgrims.
- The review methodology was robust, following a pre-registered PROSPERO protocol and PRISMA 2020 guidelines, with two independent reviewers involved in all stages of study selection, data extraction, and quality assessment to minimize bias.
- The evidence base consists solely of observational, primarily cross-sectional studies, which limits the ability to establish causality between the identified risk factors and hypertension.
- The included studies exhibited significant heterogeneity in the definitions of some exposures (e.g., dyslipidemia) and population characteristics, which impacted the consistency and certainty of some pooled estimates.
- The analysis relies on data from published studies and may be limited by potential residual confounding from variables not consistently reported in the primary articles, such as specific lifestyle factors (diet, physical activity).

## INTRODUCTION

Hypertension is a leading global public health challenge and the most significant modifiable risk factor for cardiovascular disease (CVD) and premature death worldwide [1]. The global burden of hypertension is substantial, affecting over 1.2 billion people and contributing to millions of deaths annually from complications such as ischemic heart disease, stroke, and kidney failure [1]. This burden is particularly pronounced in the context of mass gatherings, where environmental stressors, physical exertion, and psychological strain can exacerbate pre-existing chronic conditions [2].

The Hajj pilgrimage to Mecca, Saudi Arabia, is one of the largest and most demanding annual mass gatherings. It requires pilgrims to undertake a series of physically strenuous rituals in a crowded and often climatically harsh environment, posing significant health risks, especially for vulnerable individuals [3]. Indonesia consistently sends the largest national contingent of pilgrims, with over 200,000 participants each year. A unique characteristic of this cohort is its demographic profile: a large proportion are elderly, and many have been on waiting lists for decades, during which time the prevalence of non-communicable diseases (NCDs) has increased [4]. Consequently, a majority of Indonesian pilgrims are classified as high-risk (*Risti*) for health events.

Among NCDs, hypertension is the most prevalent condition and a primary contributor to morbidity and mortality among Indonesian pilgrims [5, 6]. Studies have shown that CVD is the leading cause of death during the Hajj, and uncontrolled hypertension is a key underlying factor [5]. While several primary studies from different Indonesian embarkation points have investigated local risk factors for hypertension [7-9], their findings have not yet been systematically aggregated to provide a comprehensive national picture. A quantitative synthesis is crucial to move beyond localized evidence, generate more precise and generalizable risk estimates, and provide a robust evidence base for national health policy.

This study, therefore, aims to be the first systematic review and meta-analysis to identify and quantify the primary risk factors associated with hypertension in the Indonesian Hajj pilgrim population. By synthesizing the available evidence, this study seeks to inform the development of targeted, evidence-based public health strategies, particularly focusing on the long-term health management of pilgrims during the extensive pre-departure waiting period.

### What is already known on this subject?

- Hypertension is a common and significant comorbidity among Hajj pilgrims globally, contributing to increased rates of morbidity and mortality.
- Individual observational studies in Indonesia have identified several risk factors for hypertension in specific pilgrim cohorts, such as advanced age, diabetes, and obesity.
- The demographic profile of Indonesian pilgrims includes a large proportion of elderly individuals, placing them at a high baseline risk for cardiovascular events.

### What does this study add?

- This is the first systematic review and meta-analysis to quantitatively synthesize evidence on hypertension risk factors specifically for the Indonesian Hajj pilgrim population.
- This study provides pooled risk estimates, confirming that Diabetes Mellitus, obesity, and family history are statistically significant risk factors, while the role of dyslipidemia appears inconsistent.
- The findings provide strong evidence to support a policy shift towards long-term, preventive health coaching during the multi-year Hajj waiting period, rather than focusing only on pre-departure screening.

## METHODS

This systematic review and meta-analysis was conducted and reported in accordance with the *Preferred Reporting Items for Systematic Reviews and Meta-Analyses* (PRISMA) 2020 statement [10]. The review protocol was pre-registered with the International Prospective Register of Systematic Reviews (PROSPERO), registration number: CRD420251084054.

### Eligibility Criteria

This review included analytical observational studies (cross-sectional, cohort, and case-control) that evaluated potential risk factors for hypertension. The study population was defined as prospective or current Hajj pilgrims from Indonesia. The primary outcome had to be the prevalence or incidence of hypertension, defined as a systolic blood pressure (SBP) of ≥140 mmHg and/or a diastolic blood pressure (DBP) of ≥90 mmHg, or a prior clinical diagnosis.

Studies were required to report a quantitative measure of association (e.g., Odds Ratio [OR], Prevalence Ratio [PR]) with a corresponding 95% Confidence Interval (CI). Reviews, editorials, case reports, experimental studies (RCTs), and studies on non-Indonesian pilgrims were excluded.

### Information Sources and Search Strategy

A comprehensive search was conducted in PubMed, Scopus, Google Scholar, and two national Indonesian databases (GARUDA, SINTA) for articles published up to June 2025. The full search strategy for PubMed is detailed in Supplementary Appendix 1.

### Selection Process

Two reviewers (author 1, author 2) independently screened all identified titles and abstracts using the Rayyan AI web application. Full texts of potentially eligible articles were then retrieved and assessed independently by the same two reviewers against the pre-defined eligibility criteria. Any disagreements at either stage were resolved through discussion and consensus.

### Data Collection Process and Data Items

Using a standardized and pre-piloted data extraction form, two reviewers independently extracted data from each included study. Data items included study information, participant characteristics, and outcome data. Study authors were not contacted for missing information.

### Study Risk of Bias Assessment

The methodological quality of included studies was independently assessed by two reviewers using the Joanna Briggs Institute (JBI) Critical Appraisal Checklist for Analytical Cross-Sectional Studies [11].

### Synthesis Methods

A narrative synthesis and a random-effects meta-analysis were performed using Review Manager (RevMan 5.4). Odds Ratios and Prevalence Ratios were analyzed as relative risk estimates. Heterogeneity was assessed using the I^2^ statistic. If substantial heterogeneity was detected (I^2^ > 75%), an exploration of its sources through subgroup analysis based on study location (embarkation point) was planned, provided sufficient data were available. Furthermore, a sensitivity analysis was planned by excluding studies with a high risk of bias to assess the robustness of the pooled results.

### Patient and Public Involvement

No patients or members of the public were involved in the design, conduct, reporting, or dissemination plans of this research. As this study is a systematic review of existing literature, direct patient and public involvement was not appropriate for the research methodology.

### Reporting Bias and Certainty Assessment

Publication bias was planned to be assessed using funnel plots for any analysis including 10 or more studies. The certainty of evidence was assessed using the GRADE framework [12].

## RESULTS

### Study Selection

The search yielded 128 records, from which 5 cross-sectional studies involving 286,987 pilgrims were included in the final analysis. The PRISMA flow diagram detailing the study selection process is presented in Figure 1. A list of studies excluded at the full-text stage, with reasons for exclusion, is available in Supplementary Appendix 2.

**Figure 1.**
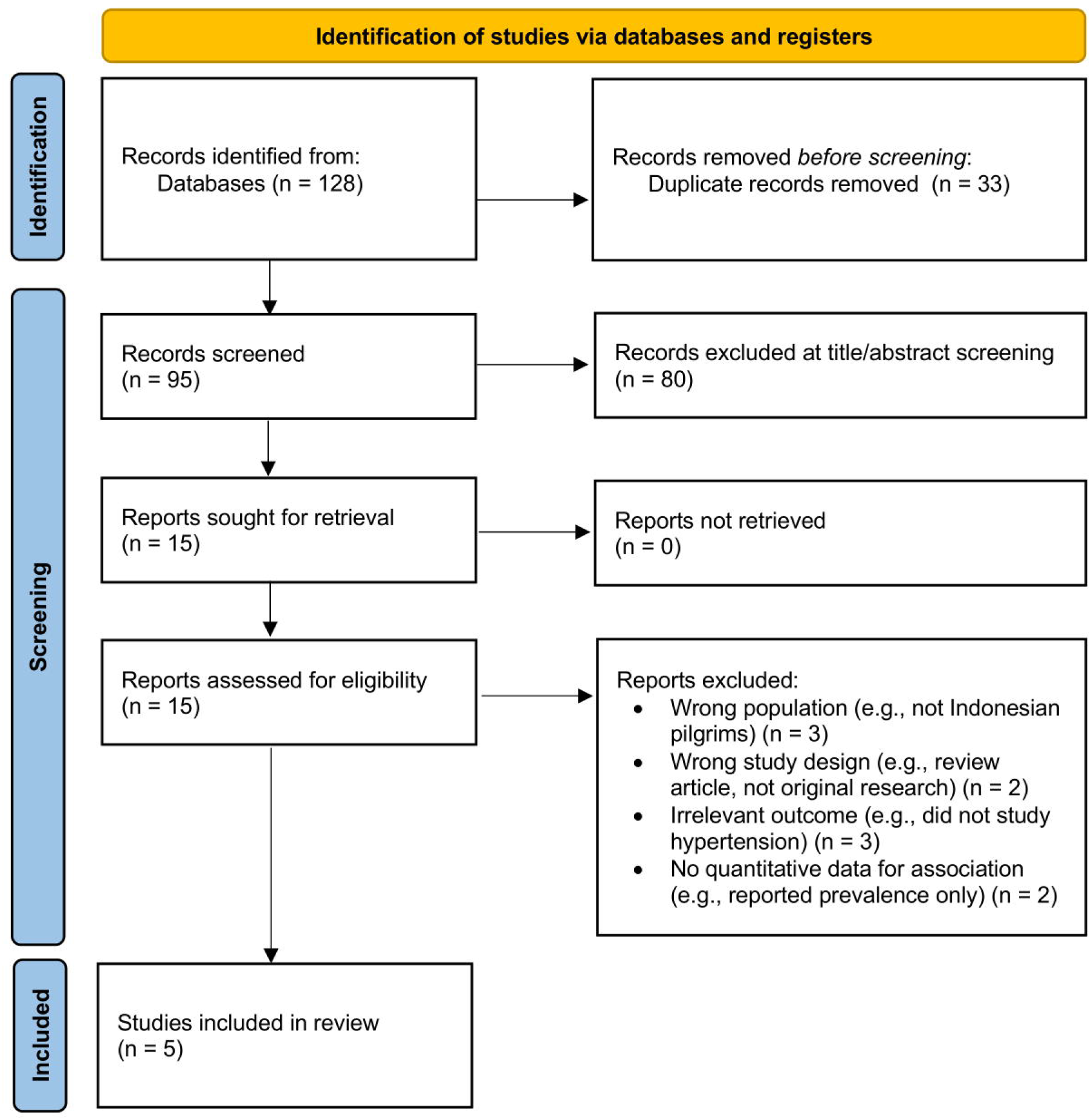
PRISMA 2020 flow diagram illustrating the study selection process.

### Study Characteristics

The five included studies were published between 2019 and 2024. All utilized data from the national Siskohatkes database. Detailed characteristics are presented in Table 1.

**Table 1.** Characteristics of Included Studies.

| Author<br>(Year) | Design | Location/<br>Embarkation | Sample (n) | Main Risk<br>Factors<br>Studied | Effect<br>Measure<br>Reported |
| --- | --- | --- | --- | --- | --- |
| Pratiwi &<br>Helda (2024)<br>[5] | Cross-sectional | DKI Jakarta | 8,014 | Age, BMI,<br>DM,<br>Dyslipidemia,<br>Family History | Prevalence<br>Ratio (PR) |
| Nani et al.<br>(2024) [6] | Cross-sectional | Banjarmasin | 5,718 | Age, BMI,<br>Central | Prevalence<br>Ratio (PR) |
|  |  |  |  | Obesity,<br>Dyslipidemia,<br>Family History |  |
| Harahap &<br>Nurwahyuni<br>(2019) [7] | Cross-sectional | Medan | 6,597 | Age, Gender,<br>Education,<br>Occupation,<br>DM, Obesity | Odds Ratio<br>(OR) |
| Liberty et al.<br>(2019) [8] | Retrospective<br>Cohort | National | 152,429 | Hyperglycemia<br>, Hypertension,<br>Central<br>Obesity | Hazard<br>Ratio (HR) |
| Ardiana &<br>Nirwana<br>(2024) [9] | Cross-sectional | National | 114,069 | Dyslipidemia<br>(HDL, LDL,<br>Triglycerides) | Odds Ratio<br>(OR) |

### Risk of Bias in Studies

The overall risk of bias in the included studies was judged to be low to moderate, with the main potential for bias arising from residual confounding due to unmeasured lifestyle factors. A detailed assessment is provided in Supplementary Figure S1.

### Results of Syntheses

Forest plots for each meta-analysis are shown in Figure 2.

**Figure 2.**
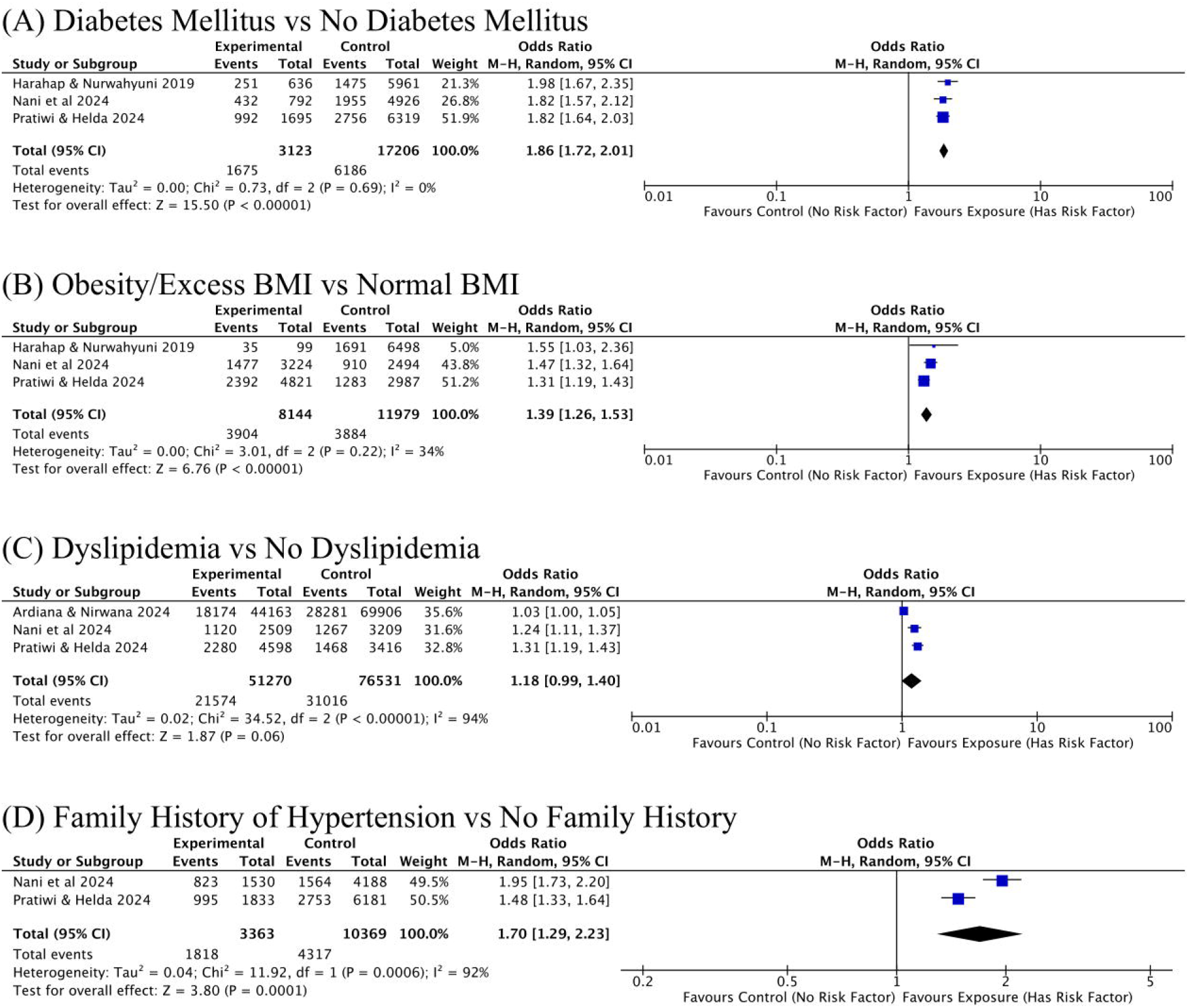
Forest plots of the association between various risk factors and hypertension. (A) Diabetes Mellitus vs No Diabetes Mellitus; (B) Obesity/Excess BMI vs Normal BMI; (C) Dyslipidemia vs No Dyslipidemia; (D) Family History of Hypertension vs No Family History.

- Diabetes Mellitus (DM): The pooled analysis of three studies showed that pilgrims with DM had a significantly higher likelihood of having hypertension (pooled OR = 1.86; 95% CI [1.72, 2.01], p < 0.00001), with no evidence of statistical heterogeneity (I^2^ = 0%).
- Obesity/Excess BMI: The meta-analysis of three studies demonstrated that obese pilgrims had a 1.39 times greater risk of hypertension (pooled OR = 1.39; 95% CI [1.26, 1.53], p < 0.00001), with moderate heterogeneity (I^2^ = 34%).
- Dyslipidemia: The pooled results of three studies indicated that dyslipidemia was not a statistically significant risk factor for hypertension (pooled OR = 1.18; 95% CI [0.99, 1.40], p = 0.06), with very high heterogeneity (I^2^ = 94%).
- Family History of Hypertension: The pooled analysis of two studies showed a 1.70 times higher risk for pilgrims with a positive family history (pooled OR = 1.70; 95% CI [1.29, 2.23], p = 0.0001), with very high heterogeneity (I^2^ = 92%).

Subgroup analysis to explore the high heterogeneity in the analyses of dyslipidemia and family history was not performed due to the limited number of included studies (fewer than three studies per potential subgroup), which would not provide sufficient power for a meaningful comparison. Similarly, a sensitivity analysis by excluding studies with a high risk of bias was not conducted, as no study was ultimately judged to have a high overall risk of bias

### Reporting Biases and Certainty of Evidence

Formal assessment of publication bias was not performed as no analysis included 10 or more studies. The certainty of the evidence for the associations, assessed using GRADE, was rated as low to moderate.

## DISCUSSION

This systematic review and meta-analysis provides the first quantitative synthesis of evidence on risk factors for hypertension among the Indonesian Hajj pilgrim population. The findings confirm that several traditional risk factors are significantly associated with hypertension, but also reveal important complexities regarding the consistency of these associations.

The most powerful and consistent finding from this meta-analysis is the strong association between Diabetes Mellitus and hypertension (pooled OR 1.86). A remarkable aspect of this result is the complete absence of statistical heterogeneity (I^2^ = 0%), indicating a highly uniform effect across diverse Indonesian pilgrim populations. This robust association is biologically plausible, as the interplay between diabetes and hypertension involves multiple shared pathophysiological pathways, including insulin resistance, activation of the renin-angiotensin-aldosterone system (RAAS), and sympathetic nervous system overactivity, all of which contribute to endothelial dysfunction and increased vascular resistance [14]. The consistency of this finding across different regions in Indonesia underscores that diabetes is a universal and critical risk factor that requires primary attention in Hajj health management.

Similarly, obesity/excess BMI was confirmed as a significant risk factor (pooled OR 1.39) with moderate heterogeneity (I^2^ = 34%). This aligns with established knowledge that adipose tissue, particularly visceral fat, functions as an active endocrine organ. It releases pro-inflammatory cytokines and adipokines that contribute to vascular inflammation, sodium retention, and increased sympathetic tone, thereby elevating peripheral resistance and blood pressure [15]. The moderate heterogeneity suggests that while the association is consistent, its magnitude may be influenced by how obesity is measured (e.g., BMI vs. central obesity) or by other population-specific characteristics.

Perhaps the most complex finding of this review relates to dyslipidemia. The pooled analysis did not find a statistically significant association between dyslipidemia and hypertension (pooled OR 1.18; 95% CI [0.99, 1.40]). This result was coupled with extremely high heterogeneity (I^2^ = 94%), which strongly suggests that it is inappropriate to conclude there is “no effect”. Rather, the evidence is highly inconsistent. This inconsistency likely stems from several sources. First, the definition of dyslipidemia varied significantly across the included studies; some used a general diagnosis, while others focused on specific lipid fractions (LDL, HDL, triglycerides), each having a different relationship with blood pressure regulation. Second, the link between lipids and blood pressure is not linear and can be heavily confounded by unmeasured variables such as diet, physical activity, and the use of lipid-lowering medications (statins), which were not accounted for in the primary studies [16]. Therefore, while dyslipidemia is a major cardiovascular risk factor, its role as a direct, independent predictor of hypertension in this specific population remains inconclusive and warrants further investigation with more standardized definitions.

A significant association was also found for a family history of hypertension (pooled OR 1.70), underscoring the interplay of genetic susceptibility and shared environmental factors.

However, this finding must be interpreted with extreme caution due to the very high heterogeneity observed (I^2^ = 92%). Such high inconsistency indicates that the magnitude of this risk factor differs substantially across the included studies. This variability is likely multifactorial. It may reflect differences in how “family history” was ascertained (e.g., accuracy of self-reporting) or true underlying differences in gene-environment interactions among diverse Indonesian sub-populations. Research has established that while genetic predisposition accounts for a significant portion of hypertension risk, its expression is heavily modulated by environmental and lifestyle factors, which can vary widely across different geographical and cultural settings within Indonesia [17].

### Strengths and Limitations

The primary strength of this review is that it is the first meta-analysis on this specific, large, and high-risk population. The use of a pre-registered protocol and adherence to PRISMA guidelines ensures methodological transparency. However, limitations include the reliance on cross-sectional studies, which prevents establishing causality, and the presence of significant heterogeneity in some analyses.

### Implications for Policy, Practice, and Future Research

The findings have profound implications. The decades-long Hajj waiting period in Indonesia presents a unique “window of opportunity” for public health intervention. The authors recommend a policy shift from short-term fitness checks to a long-term health surveillance and coaching model for all registered pilgrims, including early and periodic screening for NCDs, integrated disease management, and structured health education. Prospective cohort studies are needed to establish causality and explore the sources of heterogeneity.

## CONCLUSION

This systematic review confirms that advanced age, Diabetes Mellitus, obesity, and a family history of hypertension are significant risk factors for hypertension among Indonesian Hajj pilgrims. These findings provide a strong, evidence-based mandate for transforming the national Hajj health strategy towards a proactive, long-term, and preventive framework.

## ETHICS APPROVAL AND CONSENT TO PARTICIPATE

Ethical approval was not required for this study as it is a systematic review and meta-analysis of previously published literature. All data analyzed were sourced from publicly available studies, and no new primary data involving human participants were collected by the authors. Consent to participate was obtained by the researchers of the original primary studies included in this review.

## OTHER INFORMATION

### Support

This research received no specific grant from any funding agency.

### Competing Interests

The authors declare that they have no competing interests.

### Data Availability Statement

All data generated or analysed during this study are included in this published article and its supplementary information files.

## Notes

### Competing Interest Statement

The authors have declared no competing interest.

### Funding Statement

This study did not receive any funding

### Summary of Updates

New "Strengths and limitations" Section Added: A new summary box titled "Strengths and limitations of this study" has been added immediately after the abstract, as is standard for many BMJ journals. This section contains five bullet points focusing specifically on the study's methodology. New "Patient and Public Involvement" Section Added: A mandatory "Patient and Public Involvement" subsection has been incorporated into the Methods section to clarify that no patients were involved in the design of this systematic review. Ethics Statement Relocated: The "Ethical Considerations" statement has been moved from within the Methods section to its own dedicated section titled "ETHICS APPROVAL AND CONSENT TO PARTICIPATE" after the Conclusion, as per editorial feedback.

