## Supplementary Appendix 1 for "Risk Factors for Hypertension in Indonesian Hajj Pilgrims: A Systematic Review and Meta-Analysis"

**Supplementary Appendix 1: Full Search Strategy**

This appendix details the full electronic search strategy used for the PubMed database. This strategy was adapted for other databases, including Scopus and Google Scholar, by modifying the syntax as required.

The strategy was constructed by combining three core concepts using Boolean operators (AND, OR): (1) The Population (Indonesian Hajj Pilgrims), (2) The Outcome (Hypertension), and (3) The Exposure (Risk Factors).

**Search Strategy for PubMed Database**

| **Search** | **Query** |
|---|---|
| **#1** | "Pilgrimage"[Mesh] |
| **#2** | "Indonesia"[Mesh] |
| **#3** | Hajj[tiab] OR Hajji[tiab] OR pilgrim*[tiab] |
| **#4** | Indonesia*[tiab] OR Indonesian[tiab] |
| **#5** | #1 OR #3 |
| **#6** | #2 OR #4 |
| **#7** | #5 AND #6 |
| **#8** | "Hypertension"[Mesh] |
| **#9** | "Blood Pressure"[Mesh] |
| **#10** | hypertension[tiab] OR "high blood pressure"[tiab] OR hypertensive[tiab] |
| **#11** | #8 OR #9 OR #10 |
| **#12** | "Risk Factors"[Mesh] |
| **#13** | "Diabetes Mellitus"[Mesh] OR "Obesity"[Mesh] OR "Dyslipidemias"[Mesh] OR |

| | |
|---|---|
| | "Body Mass Index"[Mesh] OR "Hyperglycemia"[Mesh] |
| **#14** | "risk factor*"[tiab] OR determinant*[tiab] OR predictor*[tiab] OR comorbidity[tiab] OR "metabolic syndrome"[tiab] |
| **#15** | #12 OR #13 OR #14 |
| **#16** | **#7 AND #11 AND #15** |

*Note: [Mesh] is used to search for Medical Subject Headings terms, and [tiab] is used to search for keywords in the Title and Abstract. The asterisk * is a truncation character to find variations of a root word.*
