## Supplementary Appendix 1 for "Risk Factors for Hypertension in Indonesian Hajj Pilgrims: A Systematic Review and Meta-Analysis"

**Supplementary Appendix 2: List of Excluded Studies**

This appendix lists the studies that were retrieved for full-text assessment but were subsequently excluded from the review, along with the primary reason for exclusion.

| First Author (Year) | Title of Study | Reason for Exclusion |
|---|---|---|
| Naim, J. (2021) [1] | Determinants of Coronary Heart Disease Incidence among Indonesian Hajj Pilgrims Hospitalized in Saudi Arabia in 2019. | **Wrong Outcome:** The primary outcome analyzed was Coronary Heart Disease (CHD), not hypertension. Although hypertension was assessed as a predictor for CHD, it was not the dependent variable of the study. |
| Yulianto, A. (2023) [2] | Perspektif Kesehatan Matra dalam Manajemen Penyakit Tidak Menular pada Jemaah Haji: Tinjauan Literatur. | **Wrong Study Design:** The article was a narrative literature review and did not contain original research data suitable for meta-analysis. |
| Rustika, et al. (2020) [3] | Health Problems of Indonesian Hajj Pilgrims in Saudi Arabia. | **Wrong Study Design:** This article was a review abstracting three other studies, not a primary research paper. |
| Mouhtadi, R. (2023) [4] | Hubungan Kepatuhan Minum | **No Quantitative Data for** |

| | | |
|---|---|---|
| | Obat dengan Pengetahuan tentang Hipertensi pada Calon Jemaah Haji. | **Association:** The study analyzed the correlation between knowledge and medication adherence, but did not report risk ratios or odds ratios for the primary risk factors of interest (e.g., diabetes, obesity). |
| Al-Ghamdi, M. (2021) | Health profile and prevalence of chronic diseases among pilgrims attending primary health care centers in Makkah, Saudi Arabia. | **Wrong Population:** The study population consisted of a mix of international pilgrims evaluated in Saudi Arabia, not specifically Indonesian pilgrims during pre-departure screening. |
| Hakim, F.R. (2017) [5] | Hubungan Obesitas Sentral Dengan Kejadian Hipertensi Pada Jemaah Haji Kabupaten Cirebon Tahun 2017. | **No Quantitative Data for Association:** The full text was a university thesis that reported a significant p-value from a chi-square test but did not provide an Odds Ratio or Prevalence Ratio with a 95% Confidence Interval, making it |

| | | |
|---|---|---|
| | | ineligible for meta-analysis. |
| Widad, A.N. (2023) [6] | Manajemen Pelayanan Kesehatan Jemaah Haji Di Dinas Kesehatan Kota Yogyakarta pada Tahun 2022. | **No Quantitative Data:** The study was qualitative, using interviews and observation to describe health service management, and did not provide quantitative data for analysis. |
| Yezli, S., et al. (2021) | Prevalence of Diabetes and Hypertension among Hajj Pilgrims: A Systematic Review. | **Wrong Study Design:** This article is a systematic review and therefore represents secondary research, which was an exclusion criterion. |
| Ardiana, M., et al. (2023) [7] | The relationship between risk factors owned by pilgrims with hospitalization and mortality that occurs during the Hajj period. | **Wrong Outcome:** The study's outcomes were hospitalization and mortality. While it analyzed risk factors like diabetes, the dependent variable was not the incidence or prevalence of hypertension itself. |
| Purwanto, S. (2016) [8] | Indeks Rawat Inap di Arab Saudi Jemaah Haji Embarkasi | **Wrong Outcome:** The study focused on hospitalization rates |

| | | |
|---|---|---|
| | Surabaya dengan Hipertensi. | among pilgrims who already had hypertension, rather than identifying risk factors for developing hypertension. |
