## Supplementary Figure S1 for "Risk Factors for Hypertension in Indonesian Hajj Pilgrims: A Systematic Review and Meta-Analysis"

**Supplementary Figure S1: Risk of Bias Assessment of Included Studies**

This figure presents the results of the risk of bias assessment for the five included studies using the Joanna Briggs Institute (JBI) Critical Appraisal Checklist for Analytical Cross-Sectional Studies.

**A. Risk of Bias Graph: "Traffic Light Plot"**

*This plot details the judgment for each study across each of the eight JBI domains.*

| **Study** | **Q1** | **Q2** | **Q3** | **Q4** | **Q5** | **Q6** | **Q7** | **Q8** |
|---|---|---|---|---|---|---|---|---|
| Pratiwi & Helda (2024) [7] | 🟢 | 🟢 | 🟢 | 🟢 | 🟡 | 🟢 | 🟢 | 🟢 |
| Nani et al. (2024) [8] | 🟢 | 🟢 | 🟢 | 🟢 | 🟡 | 🟢 | 🟢 | 🟢 |
| Harahap & Nurwahyuni (2019) [9] | 🟢 | 🟢 | 🟢 | 🟢 | 🟡 | 🟢 | 🟢 | 🟢 |
| Liberty et al. (2019) [6] | 🟢 | 🟢 | 🟢 | 🟢 | 🟡 | 🟢 | 🟢 | 🟢 |
| Ardiana & Nirwana (2024) [13] | 🟢 | 🟢 | 🟢 | 🟢 | 🟢 | 🟢 | 🟢 | 🟢 |

**B. Risk of Bias Summary: Proportion of Studies**

*This bar chart graph summarizes the proportion of studies judged to have low, unclear, or high risk of bias across all included studies for each domain.*

| **Domain (JBI Question)** | **Low Risk (🟢)** | **Unclear Risk (🟡)** | **High Risk (🔴)** |
|---|---|---|---|
| **Q1: Clear inclusion criteria?** | 100% | 0% | 0% |
| **Q2: Subjects & setting described?** | 100% | 0% | 0% |
| **Q3: Valid exposure measurement?** | 100% | 0% | 0% |
| **Q4: Standard outcome criteria?** | 100% | 0% | 0% |
| **Q5: Confounding factors identified?** | **20%** | **80%** | **0%** |
| **Q6: Strategies to deal with confounders?** | 100% | 0% | 0% |
| **Q7: Valid outcome measurement?** | 100% | 0% | 0% |
| **Q8: Appropriate statistical analysis?** | 100% | 0% | 0% |

Figure Legend:

(A) Detailed risk of bias judgment for each included study across the eight domains of the JBI checklist. (B) Summary of risk of bias judgments presented as percentages across all included studies.

Legend: 🟢 Low risk of bias; 🟡 Unclear risk of bias; 🔴 High risk of bias.

JBI Domains:

- Q1: Inclusion criteria;
- Q2: Subject and setting description;
- Q3: Valid and reliable exposure measurement;
- Q4: Standard, objective criteria for outcome;
- Q5: Identification of confounding factors;
- Q6: Strategies to deal with confounders;
- Q7: Valid and reliable outcome measurement;
- Q8: Appropriate statistical analysis.
