## Supplementary Table S1 for "Risk Factors for Hypertension in Indonesian Hajj Pilgrims: A Systematic Review and Meta-Analysis"

**Supplementary Table S1: Summary of Findings**

Question: What are the risk factors for hypertension in Indonesian Hajj pilgrims?

Setting: Pre-departure health examinations in Indonesia.

Basis for the assumed risk (comparison group): The baseline risk of hypertension in the control group (pilgrims without the specific risk factor) is assumed to be 395 per 1,000, based on the prevalence in the non-exposed groups from the included studies.

| **Outcome** | **Assumed Risk (Control Group)** | **Corresponding Risk (Intervention Group)** | **Relative effect (95% CI)** | **No. of Participants (Studies)** | **Certainty of the evidence (GRADE)** | **Comments** |
|---|---|---|---|---|---|---|
| Hypertension in pilgrims with Diabetes Mellitus vs. without DM | 395 per 1,000 | 735 per 1,000 (679 to 794) | OR 1.86 (1.72 to 2.01) | 20,329 (3 studies) | ⊕⊕⊕O MODERATE | Downgraded one level for serious risk of bias (potential residual confounding). Not downgraded for inconsistency as $I^2$=0%. |

| | | | | | | |
|---|---|---|---|---|---|---|
| Hypertension in pilgrims with Obesity/High BMI vs. with Normal BMI | 395 per 1,000 | 502 per 1,000 (474 to 533) | OR 1.39 (1.26 to 1.53) | 20,013 (3 studies) | ⊕⊕OO LOW | Downgraded one level for serious risk of bias and one level for moderate inconsistency ($I^2$=34%). |
| Hypertension in pilgrims with Dyslipidemia vs. without Dyslipidemia | 395 per 1,000 | 448 per 1,000 (391 to 506) | OR 1.18 (0.99 to 1.40) | 125,583 (3 studies) | ⊕OOO VERY LOW | Downgraded one level for serious risk of bias, two levels for very serious inconsistency ($I^2$=94%), and the result is not statistically significant. |

| | | | | | | |
|---|---|---|---|---|---|---|
| Hypertension in pilgrims with a Family History vs. without Family History | 395 per 1,000 | 580 per 1,000 (486 to 672) | OR 1.70 (1.29 to 2.23) | 13,717 (2 studies) | ⊕OOO VERY LOW | Downgraded one level for serious risk of bias and two levels for very serious inconsistency ($I^2$=92%). |

**Explanations**

CI: Confidence Interval; OR: Odds Ratio.

**GRADE Working Group grades of evidence:**

- High certainty (⊕⊕⊕⊕): We are very confident that the true effect lies close to that of the estimate of the effect.
- Moderate certainty (⊕⊕⊕O): We are moderately confident in the effect estimate: The true effect is likely to be close to the estimate of the effect, but there is a possibility that it is substantially different.
- Low certainty (⊕⊕OO): Our confidence in the effect estimate is limited: The true effect may be substantially different from the estimate of the effect.
- Very low certainty (⊕OOO): We have very little confidence in the effect estimate: The true effect is likely to be substantially different from the estimate of effect.
